# The specter of Manaus: the risks of a rapid return to pre-pandemic conditions after COVID-19 vaccine rollout

**DOI:** 10.1101/2021.05.14.21257250

**Authors:** Debra Van Egeren, Madison Stoddard, Alexander Novokhodko, Michael Rogers, Diane Joseph-McCarthy, Bruce Zetter, Arijit Chakravarty

## Abstract

The development and deployment of several SARS-CoV-2 vaccines in a little over a year is an unprecedented achievement of modern medicine. The high levels of efficacy against transmission for some of these vaccines makes it feasible to use them to suppress SARS-CoV-2 altogether in regions with high vaccine acceptance. However, viral variants with reduced susceptibility to vaccinal and natural immunity threaten the utility of vaccines, particularly in scenarios where a return to pre-pandemic conditions occurs before the suppression of SARS-CoV-2 transmission. In this work we model the situation in the United States at present, to demonstrate how the P.1 variant of SARS-CoV-2 can cause a rebound wave of COVID-19 in a matter of months, similar to what happened in Manaus at the beginning of this year. A high burden of morbidity (and likely mortality) remains possible, even if the vaccine is partially effective against new variants and widely accepted. Our modeling suggests that variants that are already present within the population may be capable of quickly defeating the vaccines as a public health intervention, a fatal flaw in strategies that emphasize rapid reopening before achieving control of SARS-CoV-2.

## Introduction

The ongoing COVID-19 pandemic has taken a heavy toll on global health, prosperity, and stability. Within the United States, model-based estimates of mortality over the past year suggest that the pandemic is now the single deadliest event in the history of the republic^1^. Understandably, the recent deployment of several highly efficacious vaccines has led to a wave of optimism, with widespread coverage in the lay press^2–8^ heralding a return to normalcy in the coming months.

The emergence of immune-evading variants of SARS-CoV-2 darkens this otherwise rosy picture. A number of newly emerged variants have been demonstrated experimentally to be more capable of infecting cells, spreading between hosts, and/or evading natural immunity, vaccines and therapeutics, compared to the original wild-type (WT) SARS-CoV-2^9–11^. Once variants emerge within a population, they tend to expand rapidly and deterministically due to natural selection, and dominate the local viral population in a relatively short period of time^12^. These variants have in a number of cases been associated with more severe disease outbreaks, and some variants have been demonstrated to be less susceptible to certain vaccines^9,10,13,14^.

As public health authorities struggle with the problem of vaccine hesitancy, a strategy of trading away non-pharmaceutical interventions (e.g., mask wearing and social distancing) to incentivize higher vaccine uptake has emerged, both in the United States and elsewhere. A number of public health figures and media outlets have explicitly endorsed or encouraged the removal of restrictions as the vaccine rollout proceeds^2,15–17^ and public health authorities are clearly listening^18–21^. One scenario supporting this strategy is the development of herd immunity, where transmission of the virus is blocked by high levels of immunity within the population, which has been shown to be within reach given the high efficacy rates of some of the vaccines^22^. A second scenario supporting the use of this strategy is when high levels of vaccine acceptance lead to the stochastic extinction of potentially problematic variants of the virus due to drift, and drive transmission down without the need for further interventions. A case can be made that this public health outcome may in fact have been achieved successfully in some parts of the world, for example in Israel and Gibraltar^23–25^, where high rates of vaccination were followed by a gradual relaxation of restrictions. Notably, the extermination of problematic viral variants due to stochastic events may be feasible even in the absence of herd immunity, provided viral transmission is low enough^12,26^.

Unfortunately, in a situation where vaccine-evading variants are circulating widely within the population as the vaccine is deployed, evolution and infectious disease dynamics would be expected to play out quite differently. In the presence of a widely deployed vaccine, natural selection will act to enrich for the vaccine-resistant variants. Under conditions of high transmission, with a large pre-existing vaccine-resistant viral population, this can lead to a scenario where variants substantially reduce vaccine efficacy on a population level.

In this work, we examine the practical implications of the strategy of vaccinating widely and attempting a return to pre-pandemic conditions, using as our example the situation in the United States in the coming months. As of this writing, the United States has vaccinated 46% of its adult population^27^ and vaccines are expected to reach 70% of the adult population by July^28^. As 25% of the United States population consists of children, and vaccine acceptance among the 12-15 year old population (newly eligible for the vaccine) is expected to be lower than acceptance for adults^29^, one can roughly estimate that the fraction of the total population vaccinated by July would be around 50%. There are five variants of concern as defined by the CDC present at appreciable frequencies in the United States at present (see Table S1 for details): B.1.427 and B.1.429 with a transmissibility 20% greater than ancestral Wuhan strain (Wuhan-Hu-1, referred to here as “WT”); B.1.1.7 (the “United Kingdom” variant), with a transmissibility ∼60% higher than wild type^10^ and vaccine efficacy reduction of ∼10% against the Pfizer vaccine^30^; B.1.351 (the “South African” variant), with a transmissibility ∼50% higher than WT^31^ and vaccine efficacy reduction of ∼25% against the Pfizer vaccine^30^; and P.1 (the “Brazilian” variant), with a transmissibility ∼100% higher than WT^32^ and a ∼32% reduction of immunity induced by WT infection^32^. As of 4/10/2021, B.1.1.7, B.1.351 and P1 constitute ∼60%, 1% and 3.7% of all US infections^33^ (Table S1). There are other variants that have emerged recently, such as B.1.617 (the “Indian” variant) that are not yet fully characterized and may emerge as a threat during the timeline examined here, but we have not considered these.

We have used a model incorporating the dynamics of immune-evading variants to ask the question: “Do the expected levels of vaccine coverage in the US allow us to return to normal without suppressing viral transmission first?” Using a Susceptible-Infected-Recovered (S-I-R) epidemiological model with two or more competing variants with simulation conditions mirroring the current situation in the United States, we modeled the impact of vaccine-evading variants on the course of the COVID-19 pandemic in the presence of vaccines.

## Methods

We used an S-I-R model with additional infected and recovered compartments for one or more variants with different transmissibilities and conferring different immunities to other variants (Fig. 1A). This model also includes a separate population of vaccinated individuals and accounts for waning immunity over time for both vaccinated and previously infected individuals (see Supplemental Methods for details). Specifically, we assumed that vaccinated individuals could not be infected by WT virus but could be infected by variants. However, vaccination and previous infections with other variants confers partial protection against variant infection. Initially, we assumed that a single variant (B.1.1.7, P.1, or B.1.351) was present in the population at the frequency estimated in the US population on 4/10/21. We then extended the S-I-R model shown in Fig. 1A to include three competing variants simultaneously, and we used this extended model to simulate the case in which all three variants were present to investigate possible clonal interference between the variants. A more extensive description of the methods is given in the Supplemental Methods, along with parameter values used in the simulations (Tables S1 and S2).

**Figure 1.**
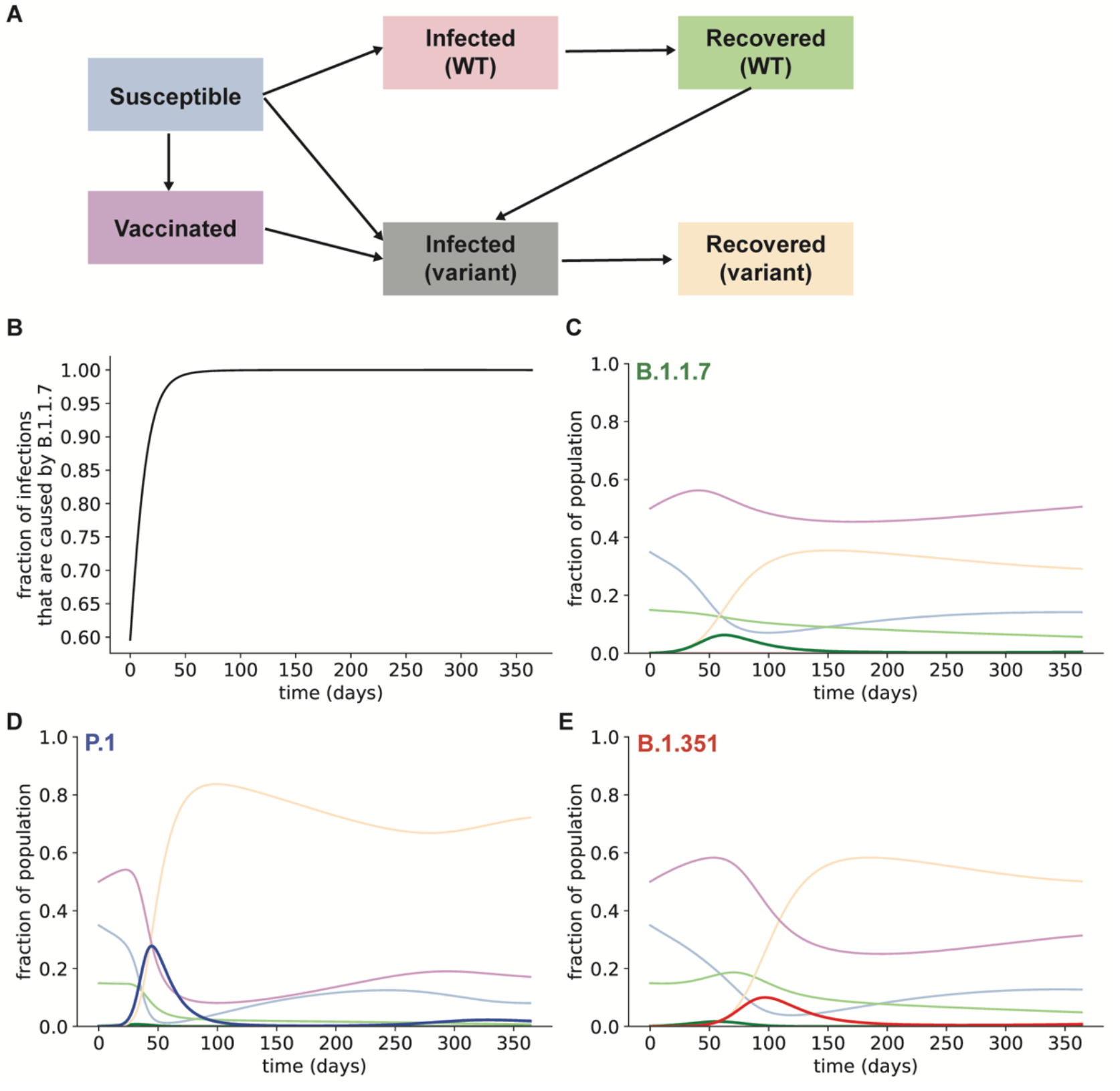
Existing SARS-CoV-2 variants are likely to cause a surge of infections in the US after reopening. **A**. Structure of S-I-R model. Susceptible individuals can be infected with WT or variant virus, while individuals with immunity to the WT virus can only be infected by variants. Individuals can also lose protective immunity and rejoin the susceptible population (not shown in this schematic). **B**. Fraction of active infections caused by the B.1.1.7 variant, assuming no additional variants are present in the populations. **C**. Simulated frequencies of infections caused by the B.1.1.7 variant (green), assuming it is the only variant present in the population. **D**. Simulated frequencies of infections caused by the P.1 variant (blue), assuming all other infections are caused by the B.1.1.7 variant (green). **E**. Simulated frequencies of infections caused by the B.1.351 variant (red), assuming all other infections are caused by the B.1.1.7 variant (green). For **C-E**, frequencies of other compartments are plotted in the colors used in the schematic in **A**.

## Results

We simulated the spread of three existing SARS-CoV-2 variants (B.1.1.7, P.1, and B.1.351) in the US if the contact rate (the maximum rate at which infected individuals can spread the infection, as dictated by their interactions with others) is allowed to return to pre-pandemic levels. Assuming a moderate level of vaccine coverage similar to current conditions in the US (50% of the population are initially fully vaccinated), we found that the B.1.1.7 variant will quickly expand and become responsible for nearly all infections (Fig. 1B-C), as it did in the UK^10^. However, the other two variants, which are more capable of infecting individuals who were vaccinated or have been previously infected with WT virus, will cause a spike of infections months after the return to normal contact levels. If only P.1 and B.1.1.7 are present in the population, P.1 will cause a wave of infections affecting >80% of the population, including those who have been vaccinated (Fig. 1D). Without P.1, the B.1.351 variant will still cause a wave of infections, although it will occur later and affect fewer individuals (Fig. 1E).

Assuming all three variants are initially present in the population (at the frequencies estimated by the CDC as of 4/10/21), if a full return to pre-pandemic conditions is attempted after vaccination, P.1 will become the dominant strain in the US within a matter of months (Fig. 2). This surge in variant infections is predicted to lead to hundreds of millions of COVID-19 cases, as well as millions of COVID-19 fatalities (Table 1). The predicted fatalities shown in Table 1 were calculated using a base infection fatality rate of 0.68%^34^ (Methods). We also assumed that vaccination results in a 70% reduction in risk of death given infection, which was observed for the Pfizer vaccine for wild-type SARS-CoV-2^35^. Variant infections (notably, P.1) will be responsible for the vast majority of cases and will increase the total infection burden approximately 100-fold (Table 1). These results suggest that, in the presence of vaccine-evading strains, a return to pre-pandemic levels of contact following vaccination is likely to result in unsustainable mortality and morbidity burdens.

**Table 1.** A return to pre-pandemic levels of mobility following vaccination is made infeasible by the presence of vaccine-evading variants. Predicted SARS-CoV-2 infection and mortality burdens over one year upon a return to pre-pandemic levels conditions (*R*_0_ = 3.32), in the United States. The predicted fatalities were calculated using a base infection fatality rate of 0.68% and assuming a 70% reduction in risk of death given infection for vaccinated individuals.

| Scenario | Total infections (millions) | Total fatalities (millions) |
| --- | --- | --- |
| Only WT | 3.5 | 0.02 |
| WT + B.1.1.7 | 148.3 | 0.9 |
| B.1.1.7 + B.1.351 | 269.7 | 1.5 |
| B.1.1.7 + P.1 | 370.1 | 2.1 |
| WT + all 3 variants | 373.7 | 2.1 |

**Figure 2.**
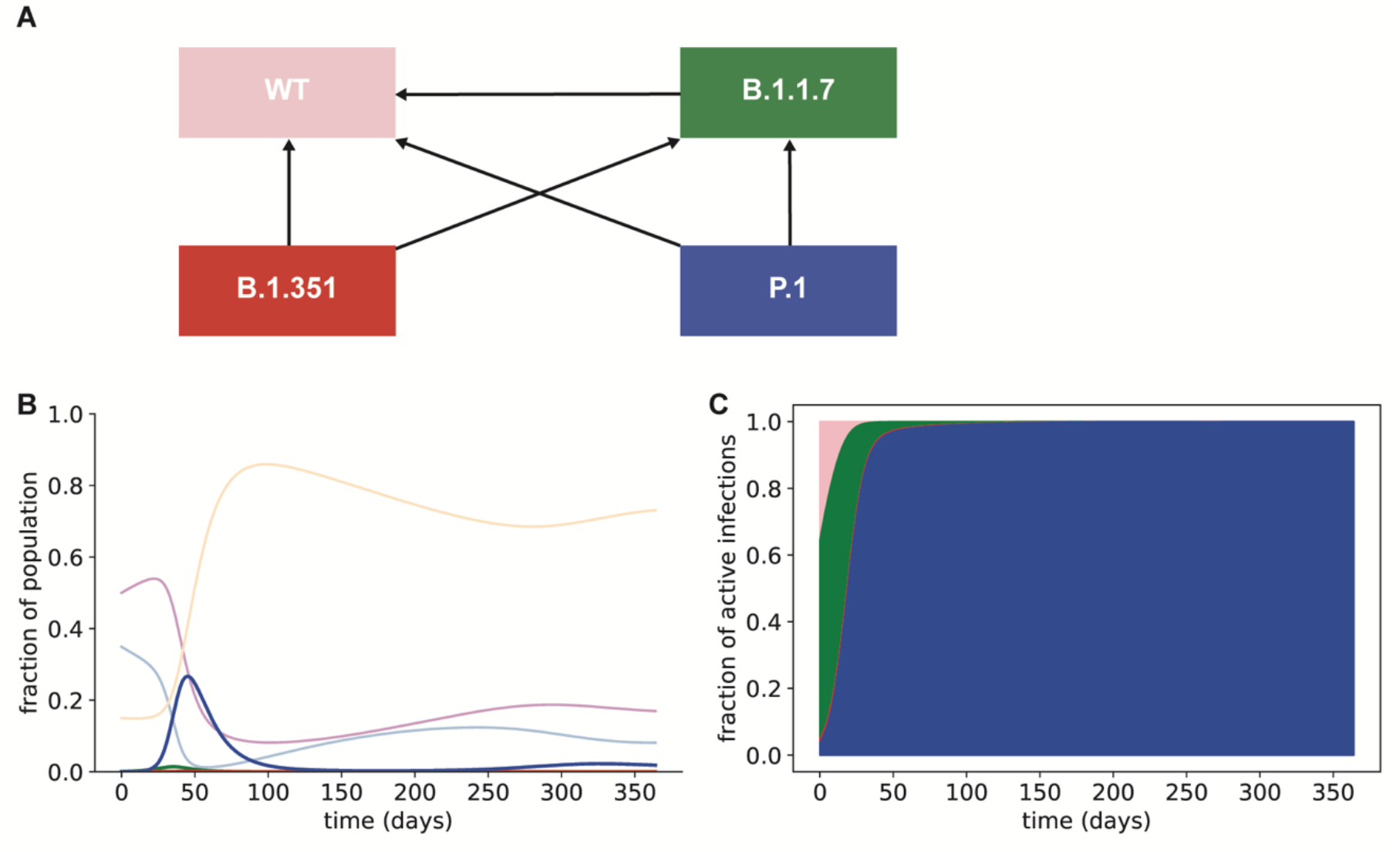
The P.1 variant is likely to outcompete other existing variants and drive a surge of infections even in a population with high WT immunity. **A**. Schematic showing cross immunity between variants used in our simulations. Arrows show which variants can infect individuals with immunity to other variants. For example, the arrow from B.1.351 to WT indicates that B.1.351 can infect individuals with immunity to the WT virus acquired by prior infection or vaccination. **B**. Fraction of individuals infected with SARS-CoV-2 variants (colors as in **A**), simulated over 1 year. Susceptible (light blue), vaccinated (light purple), and recovered from SARS-CoV-2 infection with any viral genotype (orange) fractions are also plotted. **C**. Fraction of active infections caused by each existing SARS-CoV-2 variant over the first year (colors as in the schematic in **A**).

Continuing to suppress the overall SARS-CoV-2 reproduction number (*R*_T_) with non-pharmaceutical interventions (e.g., mask wearing, social distancing) can delay the surge of variant infections and reduce overall infection burden (Fig. 3). The basic reproductive number *R*_0_ in Wuhan under normal contact levels for the WT strain was estimated to be 3.32^36^, though estimates of this value have been as high as 5.7^37^. Reducing this rate to approximately 1 for WT SARS-CoV-2 substantially suppresses the spread of all three variants, limiting the number of total infections over the next year. Therefore, our results indicate that non-pharmaceutical interventions will remain important in controlling the COVID-19 pandemic, particularly as more transmissible and immune evading variants become more frequent in the US.

**Figure 3.**
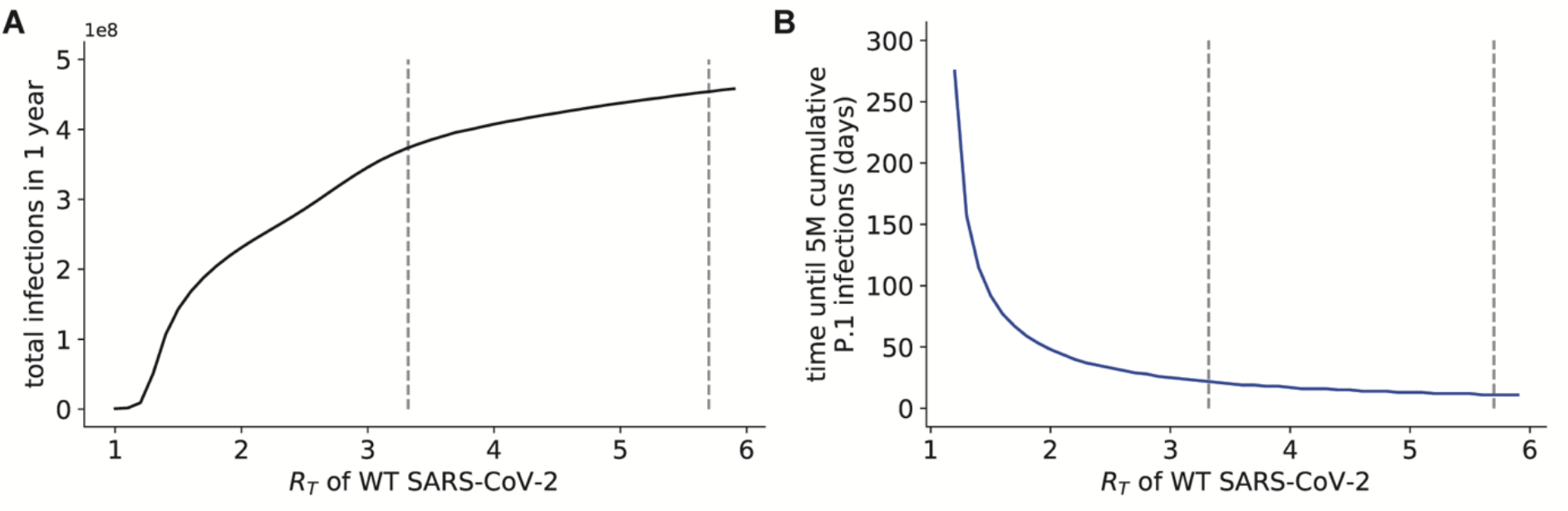
Reopening with an increased infection contact rate will lead to high total infection burden and rapid transmission of the P.1 variant in the US. **A**. Total infection burden for all variants over different contact rates corresponding to different *R*_T_ values for WT SARS-CoV-2. **B**. Length of time from the start of the simulation until 5 million individuals have been infected with the P.1 variant, given different contact rates corresponding to different *R*_T_ values for WT SARS-CoV-2. Vertical dashed lines in both panels correspond to *R*_0_ values measured at the beginning of the pandemic (3.32 and 5.7), before infection control policies were implemented.

## Discussion

In this work we demonstrate that a return to pre-pandemic conditions following modestly high levels of vaccination will efficiently select for pre-existing vaccine-evading viral variants within the population, causing a high level of infection and potentially death. While these results paint an alarming picture, they are not intended as a specific prediction of the future. Rather, we sought to illustrate a scenario in which the pandemic may-counterintuitively-persist despite high rates of vaccination after reopening. In particular, we focused on the consequences of a return to pre-pandemic conditions after moderate to high levels of vaccination, while vaccine-evading variants are still circulating in the population. Our findings suggest that this strategy is infeasible.

Our work has a few important limitations. First, the simulations of the United States shown here are examples based on the currently available estimates for viral parameters such as transmissibility and vaccine evasion. Those estimates are preliminary and may change as more information becomes available. The results here depend heavily on the estimates for transmissibility for the immune-evading strains (P.1 and B.1.351), and these estimates are vulnerable to experimental error^38^. Thus, while our work suggests, for example, that P.1 is likely to pose a public health threat in the coming months within the United States if contact rates return to pre-pandemic levels, uncertainty around viral parameters for P.1 prevents us from making a definitive prediction. Second, a number of key parameter values in our system are currently unknown: for example, vaccine impact on the transmission of viral variants, the degree of cross-protection against reinfection conferred by infection with viral variants, and the variant-specific risk of death if infected after vaccination. The estimates used in our simulations were chosen to be conservative with respect to these parameters, but emerging experimental data may change these results. Third, we do not consider the generation of new, possibly more transmissible or immune-evading variants. New variants, such as B.1.617, which recently appeared in India, could overtake existing variants and possibly lead to more infections. Finally, our modeling assumes that contact rates are fixed over the entire time period, and in practice these rates are strongly impacted by current events^39,40^, as populations experiencing outbreaks of COVID-19 tend to spontaneously adopt non-pharmaceutical interventions (mask-wearing, reduction in levels of mobility, social distancing) when hospitalization and death rates climb. As a result, the findings here represent a worst-case scenario, in the complete absence of rational behavior within the impacted population.

Nevertheless, or perhaps precisely because they represent a worst-case scenario, the simulations here provide some key lessons for the next phase of the pandemic. Crucially, our work points to the importance of keeping non-pharmaceutical interventions in place to suppress transmission of SARS-CoV-2 both before and for a period after vaccine rollout. The contact rate has a large impact on both the timing and the magnitude of subsequent waves of vaccine-resistant variants, and if the contact rate is sufficiently low, such waves will not materialize at all. For example, leveraging the ability to reduce indoor transmission during the summer in the US may reduce the impact of the variants over the next few months. Thus, combining non-pharmaceutical interventions with vaccine- and naturally-induced immunity, even in the absence of herd immunity, may be a practical path forward for achieving SARS-CoV-2 suppression. Our work also suggests a metric that will yield critical information in the coming months-the absolute case counts for different variants of concern. An increase in absolute numbers of variants such as P.1 in the United States during the reopening, particularly after the bulk of the population is vaccinated, will be an early warning sign of a public health crisis.

Our results also point out two significant misconceptions in the commonly held view of the SARS-CoV-2 variants at present-the belief that the “vaccines will still work”^41–43^ and the idea that the vaccines can “win the race” against the variants^44,45^. Our work shows that even relatively small reductions in immune protection (such as those reported for P.1 and WT reinfection) can have catastrophic consequences in the face of high contact and infection rates, and that high levels of pre-existing immunity will only serve to select efficiently for immune-evading variants. Thus, the vaccines may *not* work well enough prevent mortality on the population level, and it is *not* a race. If restrictions are eased too rapidly, leading to high contact rates and high levels of viral transmission, immune-evading variants of SARS-CoV-2 are likely to expand efficiently and deterministically, leading to further waves of COVID-19. There is evidence that this scenario has already come to pass elsewhere in the world-in Manaus, after the first wave of infections in late 2020^14,46,47^, and in Chile and the Seychelles as of this writing^48,49^. Our models suggest that high levels of ongoing contact in the presence of the virus will rapidly lead to disease resurgence, and this may be an intrinsic feature of the current pandemic that is at once unintuitive and deadly. Those who forget the recent past may be condemned to repeat it.

## Data Availability

No new experimental or clinical data were collected. Only simulation results are presented in the paper.

