## Supplemental Methods and Tables for "The specter of Manaus: the risks of a rapid return to pre-pandemic conditions after COVID-19 vaccine rollout"

##### Detailed S-I-R model description

To investigate SARS-CoV-2 infection dynamics and evolution after vaccine deployment, we built a deterministic susceptible-infected-recovered (S-I-R) epidemiological model that explicitly accounts for the spread of existing variants. The simplest S-I-R model we used has two competing viral genotypes circulating in the population: wild-type (WT) and variant SARS-CoV-2 (Fig. 1A). This model has separate infected and recovered compartments for individuals with WT and variant infections. Importantly, vaccination and infection with WT virus completely protects individuals from infection with WT virus, but only provides partial protection from variant infection. This overestimates vaccine efficacy against WT SARS-CoV-2, as the most effective vaccines (the mRNA vaccines deployed widely in the US) have been shown to be approximately 95% effective against infection [1].

The model equations are

$$
\begin{aligned}
\frac{dS}{dt} &= \rho(R + R_m + V) - (\beta I + \beta\alpha_m I_m + z)S \\
\frac{dV}{dt} &= zS - (\rho + \beta(1-c)\alpha_m I_m)V \\
\frac{dI}{dt} &= \beta S I - \gamma I \\
\frac{dI_m}{dt} &= (S + (1-c)(R+V))\beta\alpha_m I - \gamma I_m \\
\frac{dR}{dt} &= \gamma I - (\rho + \beta\alpha_m(1-c)I)R \\
\frac{dR_m}{dt} &= \gamma I_m - \rho R_m
\end{aligned}
$$

where $S$ and $V$ correspond to the fractions of the population who are susceptible and vaccinated, respectively, with susceptible individuals receiving the vaccine at rate $z$. $I_w$ and $I_m$ represent the fraction of individuals who are currently infected with WT or variant SARS-CoV-2, respectively. These infected individuals eventually recover at rate $\gamma$ and enter the recovered compartments, $R_w$ and $R_m$. Susceptible individuals are infected with WT virus at a rate proportional to the contact rate $\beta$. The variant infects susceptible individuals with an effective contact rate $\beta\alpha_m$, where $\alpha_m$ is the variant's transmission advantage relative to WT. Note that there is a relationship between the contact rate $\beta$, the recovery period length, and the basic reproductive number $R_0$ of the WT virus; namely, $\beta = \gamma R_0$. Vaccinated individuals and those who have recovered from WT infection are partially protected from variant infection and are infected at rate proportional to $\beta\alpha_m(1-c)$, where $c < 1$ is the degree of protection from variant infection conferred by WT immunity. Individuals who have recovered from the variant infection cannot be reinfected with either the WT or variant virus. Both vaccinal and natural immunity are lost at rate $\rho$.

This S-I-R modeling framework was expanded to include multiple competing SARS-CoV-2 variants. In the more general case, there are $n$ SARS-CoV-2 variants competing against each other and the WT virus. There are in total $n+1$ infected and $n+1$ recovered compartments, each corresponding to a specific viral genotype. Defining $\vec{I}$ and $\vec{R}$ as the vectors of frequencies of individuals infected and recovered from each viral genotype, we have

$$\frac{dS}{dt} = \rho \sum R_i - zS - (\vec{I} \cdot \mathbf{B}_S)S$$
$$\frac{dV}{dt} = zS - \rho V - (\vec{I} \cdot \mathbf{B}_V)V$$
$$\frac{d\vec{I}}{dt} = (\mathbf{B}^T \mathbf{A}) \odot \vec{I} - \gamma \vec{I}$$
$$\frac{d\vec{R}}{dt} = \gamma \vec{I} - \rho \vec{R} - \mathbf{B}_R \vec{R}^T \odot \vec{I}$$

where $\odot$ denotes element-wise multiplication. The contact rate matrix $\mathbf{B}$ describes the transmissibilities and cross infection potentials of the variants in the simulation, and has $n+3$ rows corresponding to each of the non-infected compartments ($S$, $V$, and $\vec{R}$) and $n+1$ columns corresponding to the infected compartments. Each entry $b_{i,j}$ denotes the rate at which infected compartment $j$ can infect individuals in compartment $i$. More specifically,

$$\mathbf{B} = \beta \begin{pmatrix} 1 & \alpha_1 & \dots & \alpha_n \\ 0 & \alpha_1 c_{0,1} & \dots & \alpha_n c_{0,n} \\ 0 & \alpha_1 c_{0,1} & \dots & \alpha_n c_{0,n} \\ c_{1,0} & \alpha_1 c_{1,1} & \dots & \alpha_n c_{1,n} \\ \vdots & \vdots & \ddots & \vdots \\ c_{n,0} & \alpha_1 c_{n,1} & \dots & \alpha_n c_{n,n} \end{pmatrix}$$

where $\alpha_i$ is the transmission advantage of variant $i$ and $c_{i,j}$ is the cross immunity to infection with variant $j$ conferred by previous infection with variant $i$, where variant 0 is WT. The first row (also defined as $\mathbf{B}_S$) corresponds to the rates at which the WT and variants infect susceptible individuals. Similarly, the second row (also defined as $\mathbf{B}_V$) corresponds to the rates at which the WT and variants infect vaccinated individuals. The remaining rows of this matrix are defined as $\mathbf{B}_R$, and corresponds to the rates at which the WT and variants infect recovered individuals. The cross immunity relationships shown in Fig. 2A were used to populate the entries of $\mathbf{B}$. The column vector $\mathbf{A}$ has entries $a_1 = S$, $a_2 = V$, and $a_{i+2} = R_i$ for $i = 0, \dots, n$.

To simulate both versions of the model, the above systems of differential equations were numerically solved using `scipy.integrate` (version 1.4.1) in Python. Parameter values, initial conditions, and references used for the simulations are provided in Tables S1 and S2.

#### Estimation of total fatalities due to COVID-19

The total number of deaths due to COVID-19 during the first year after the starting point of the simulations was estimated by using the simulation results to calculate the number of COVID-19 cases occurring in previously unexposed or previously-infected/vaccinated individuals separately. Individuals who were not previously exposed to SARS-CoV-2 were assumed to have an infection fatality rate (IFR) of 0.68% [2]. We estimated the IFR for vaccinated/immune individuals relative to unexposed individuals as $\frac{1-\mathrm{IRR}_{\mathrm{vac}}}{1-\mathrm{IRR}_{\mathrm{unvac}}}$, where IRR is the infection rate ratio, as measured in [3]. This corresponds to an IFR for vaccinated and previously-infected individuals of 0.48%.

### Supplemental Tables

**Table S1. Parameter values for transmissibility and cross immunity for SARS-CoV-2 variants.**

| Variant | Transmissiblity relative to WT | Trans. reference | Vaccine or WT immune efficacy | Immunity reference | % of US infections as of 4/10/21 |
|---|---|---|---|---|---|
| B.1.1.7 | 1.59 | [4] | 89.5% | [5] | 59.6% |
| B.1.351 | 1.5 | [6] | 75.0% | [5] | 1.0% |
| P.1 | 2.0 | [7] | 68.0% | [7] | 3.7% |

All variant frequency data was taken from the CDC COVID Data Tracker [8].

**Table S2. Other parameter values used to simulate SARS-CoV-2 spread.**

| Parameter | Value | Reference |
|---|---|---|
| Initial fraction of population vaccinated | 50% | [9] |
| Daily vaccine uptake rate (% susceptibles vaccinated/day) | 1% | [9] (upper bound) |
| Initial fraction of population previously infected | 30% | [10] |
| Initial fraction of population currently infected | 0.13% | $\frac{\text{(current case count)(mean infection duration)}}{\text{total US population}}$ |
| Mean infection duration | 10 days | [11] |
| Mean duration of immunity | 18 months | [12] (upper bound) |
| $R_0$ of WT SARS-CoV-2 | 3.32 | [13] |
| Total population size | 300 million | approximate size of US population |
